# Predictive modeling stroke prediction using routine clinical and socio-demographic data in Uganda

**DOI:** 10.64898/2026.10.19.26365927

**Authors:** Derrick Mugisha, Benjamin Watyaba, Haulat Kiiza, David Wagaba, Happy Unith, Shamim Nabidda, Samuel Okello, Joan N. Kalyango, Rehema Baguma, Martin Kaddu-Mukasa, Jane Nakibuuka

## Abstract

Stroke is a leading cause of mortality and disability in Uganda, with a prevalence of approximately 2.5% and a 25-30% 30-day mortality rate, exacerbated by limited access to predictive tools and advanced diagnostics. Machine learning (ML) offers potential for early stroke risk stratification, but its application in resource-constrained settings like Uganda requires tailored models and rigorous performance evaluation, as well as assessment of misclassification patterns. This study aimed to develop and internally validate an ML-based predictive model for stroke using clinical and socio-demographic characteristics among patients at Mulago National Referral Hospital (MNRH) and residents of Wakiso District, Uganda, identify key predictors and assess misclassification patterns to enhance targeted prevention.

A retrospective case-control study utilized secondary data from 1,016 participants (254 stroke cases, 762 controls) across three prior studies conducted between 2006 and 2015. Eight ML algorithms were evaluated using 5-fold cross-validation, with performance metrics including accuracy, sensitivity, positive predictive value, F1-score, and area under receiver operator characteristics curve (AU-ROC). McNemar’s test compared model classification performance of the top 2 models. Feature importance identified key predictors, and misclassification patterns were analyzed using classification matrix and line graphs.

The Random Forest (RF) model achieved the highest performance (95.18% accuracy, 90.10% F1-score, 97% AU-ROC). McNemar’s test confirmed RF’s superior classification (p=0.01). Age, systolic blood pressure, total cholesterol, and waist-hip ratio were the top drivers of RF stroke predictions. RF misclassified 8.3% of stroke cases and 3.3% of non-stroke cases, due to overlaps in risk profiles (e.g., lower cholesterol, systolic blood pressure and age in false negatives).

The RF model demonstrates strong potential for stroke risk prediction in Uganda, leveraging routine data to prioritize high-risk individuals. Misclassification analysis highlights areas for model refinement, supporting its potential for integration into clinical workflows for early intervention and stroke prevention in resource-limited settings. Future external validation across diverse populations is recommended.

## Background

Stroke is a major global health challenge, causing approximately 15 million cases annually, with 5 million deaths and 5 million permanent disabilities (1). It ranks as the third leading cause of death, accounting for 11% of global mortality (2). The vast majority of the global burden occurs in low- and middle-income countries where nearly 86% of stroke deaths are reported (3) With a prevalence of ∼2.5% and a 25-30% 30-day mortality rate, stroke contributes significantly to hospital admissions and long-term disability (4). Strokes are primarily classified as ischemic, hemorrhagic, or transient ischemic attacks (TIAs). Hemorrhagic strokes occur when brain blood vessels rupture, causing bleeding that can lead to severe brain damage, while TIAs, or “mini-strokes,” are temporary blockages in brain blood flow. Ischemic stroke accounts for approximately 62-78% of all incident strokes in low-and middle income countries (3–5), caused by clots or fatty deposits obstructing blood flow to the brain, often resulting in severe disability or death (6). The impact of stroke is profound, leading to long-term disability, reduced quality of life, and significant emotional and financial burdens for families and health systems, particularly in low-resource settings (1,7). Approximately 80% of strokes are preventable through early detection and management of risk factors, yet such strategies are underutilized in Uganda due to inadequate diagnostic access (4,8). ML offers a promising approach to enhance stroke risk prediction by analyzing complex patterns in clinical and socio-demographic data. Models like RF and logistic regression have shown success in identifying high-risk individuals, by examining data patterns that may not be evident through traditional analysis (9,10). Predictive tools like QStroke have demonstrated effective risk stratification in high-income settings, but their application in Uganda is limited by lack of validated local models, and resource constraints (11). Understanding model limitations, particularly misclassification patterns, is critical for practical implementation. Misclassified cases where stroke patients are predicted as non-stroke or vice versa can delay interventions or overburden healthcare systems, a significant concern in Uganda’s high-mortality context. Analyzing these patterns helps identify why errors occur, such as overlaps in risk factor profiles or unmeasured variables, and informs model calibration to optimize sensitivity and specificity (12). This study aimed to develop and validate a machine learning-based predictive model for stroke risk using clinical and socio-demographic characteristics among patients at MNRH and residents of Wakiso District, Uganda. By identifying key predictors, evaluating model accuracy, and assessing misclassification patterns and performance limitations, the study sought to deliver a practical tool for early stroke risk stratification, tailored to Uganda’s resource-limited healthcare system, to support targeted prevention and reduce stroke burden.

## Methods

### Study Design and Setting

This retrospective unmatched case-control study utilized secondary data from three prior studies conducted between 2006 and 2015 at MNRH, Uganda’s largest public hospital, and Wakiso District, the most populous and rapidly urbanizing region near Kampala. Mulago handles ∼1,000 stroke cases annually, while Wakiso provides a diverse community sample for controls.

### Data Sources

Data were sourced and obtained on 7^th^ March, 2025 from:

1. A non-randomized trial (2014-2015) of 254 stroke patients at Mulago (13).
2. A descriptive study (2006) of 85 stroke patients at Mulago (14).
3. A population survey (2012-2013) of 5,420 Wakiso residents without stroke history (15).

### Data Collection and Management

Data were extracted using a pre-tested abstraction tool from paper-based (cases) and electronic (controls) records into an EpiData 4.6 database. Double entry ensured accuracy, and anonymized identifiers protected confidentiality. Data were exported to Python Jupyter Notebook 3.9 for analysis.

### Study Population

The study included 1,016 participants (254 stroke cases, 762 controls). Cases were patients aged ≥18 years with confirmed stroke (via plain CT scan) within 7 days of symptom onset, admitted to Mulago hospital. Controls were Wakiso residents aged ≥18 years with no stroke history. Records with missing data were excluded.

### Sample Size and Sampling

Sample size was calculated using the Events Per Variable (EPV) method (16) (10 EPV for 26 predictors), requiring 260 cases and 780 controls (1:3 ratio). All 254 eligible cases from the 2014-2015 study were included, supplemented by six randomly selected cases from the 2006 study using NumPy’s random number generator. Controls were systematically sampled (every 6th record) from the Wakiso survey.

### Variables

The dependent variable was stroke diagnosis, defined per WHO criteria as sudden focal neurological deficits sometimes global, lasting >24 hours or causing death, with vascular origin. Predictors included:

Clinical: History of hypertension, systolic/diastolic blood pressure, total cholesterol, High Density Lipoprotein cholesterol (HDL), Low Density Lipoprotein(LDL), triglycerides, waist-hip ratio (WHR), diabetes mellitus (DM) history, fasting blood sugar, HIV sero-status.

Socio-Demographic: Age, sex.

Lifestyle: Smoking history, frequency of smoking, alcohol consumption, harmful alcohol use (via AUDIT).

### Data Analysis

Analysis used Python Jupyter notebook 3 (pandas, scikit-learn, numpy, matplotlib) with a significance level of α=0.05.

**Preprocessing:** Continuous variables were summarized with medians/IQR due to non-normal distributions (Shapiro-Wilk test). Multi-collinearity was assessed via Variance Inflation Factor (VIF<10) and Pearson’s correlation (<0.7). Recursive Feature Elimination with Cross-Validation (RFECV) selected optimal predictors. Continuous variables were standardized (mean=0, SD=1), and categorical variables were one-hot encoded. Synthetic Minority Oversampling Technique (SMOTE) addressed class imbalance.

**Model Development:** Eight ML algorithms (RF, AdaBoost, Gradient Boosting, Logistic Regression, SVM, Decision Tree, KNN, Naïve Bayes) were evaluated using 5-fold cross-validation.

**Model Validation:** Performance metrics included accuracy, sensitivity, precision, F1-score, and AU-ROC. McNemar’s test compared the top two models.

Model Interpretation: Feature importance (Gini index) identified key predictors.

Misclassification Analysis: Patterns were assessed via a classification matrix and line graph.

## Results

### Description of Clinical and Sociodemographic Characteristics of study Participants (n=1016)

The cases and controls were significantly different on many characteristics. The cases had higher proportions of males (40.4% versus 26.9%), history of hypertension (58.5% versus 5.4%), history of DM (8.5% versus 1.2%), and harmful alcohol use (12.7% versus 2.8%). In addition, cases compared to controls had higher proportions of smoking history (6.5% versus 5.3%) but lower proportions of HIV positive status (3.5% versus 9.5%). The cases were also older (median 61 years versus 30) and had higher median levels for most clinical measurements except LDL (p=0.069) and triglycerides (p=0.643). Clinical and sociodemographic characteristics are summarized in Table 1.

**Table 1:** Clinical and socio-demographic characteristics of stroke cases (N=254) and non-stroke controls (N=762).

| Characteristic | Cases (Stroke),<br>n(%) | Controls (Non-Stroke),<br>n(%) | $\chi^2$<br>Statistic | P value |
| --- | --- | --- | --- | --- |
| **Sex, n (%)** |  |  | 17.58 |  |
| Male | 103(40.5) | 201(26.4) |  | <0.001 |
| Female | 151(59.5) | 561(73.6) |  |  |
| **History of Hypertension, n (%)** |  |  | 356.02 |  |
| Yes | 150(59.1) | 41(5.4) |  | <0.001 |
| No | 104(40.9) | 721(94.6) |  |  |
| **History of Diabetes mellitus, n** |  |  | 33.55 |  |
| **(%)** | 22(8.7) | 9(1.2) |  | <0.001 |
| Yes | 232(91.3) | 753(98.8) |  |  |
| No |  |  |  |  |
| **History of Alcohol Use, n (%)** |  |  | 1.02 |  |
| Yes | 77(30.3) | 204(26.8) |  | 0.270 |
| No | 177(69.7) | 558(73.2) |  |  |
| **Harmful Alcohol Use, n (%)** |  |  | 33.10 |  |
| Yes | 31(12.2) | 21(2.8) |  | <0.001 |
| No | 223(87.8) | 741(97.2) |  |  |
| **History of Smoking** |  |  | 14.17 |  |
| Current Smoker | 0(0.0) | 40(5.2) |  | <0.001 |
| Former Smoker | 16(6.3) | 9(1.2) |  |  |
| Non-Smoker | 238(93.7) | 683(89.6) |  |  |
| **HIV, n (%)** |  |  | 8.27 |  |
| Positive | 9(3.5) | 72(9.5) |  | 0.007 |
| Negative | 245(94.5) | 690(90.5) |  |  |

|  | **Median (IQR)** | **Median (IQR)** | U |  |
| --- | --- | --- | --- | --- |
|  |  |  | Statistic |  |
| Age | 60(50-75) | 30(24-40) | 18147.00 | <0.001 |
| Fasting Blood Sugar | 6.2(5.4-7.8) | 5.1(1.9-5.6) | 41110.50 | <0.001 |
| Systolic Blood Pressure | 160 (140-180) | 120(111-132) | 31786.00 | <0.001 |
| Diastolic Blood Pressure | 95 (84-110) | 77(71-84) | 40356.50 | <0.001 |
| Pulse Rate | 81 (72-92) | 79(71-87) | 84853.50 | 0.003 |
| Waist Hip Ratio | 0.98 (0.89-1.23) | 0.82(0.78-0.87) | 25375.00 | <0.001 |
| Total Cholesterol | 5.6 (4.5-6.8) | 2.9(2.2-4.2) | 35599.50 | <0.001 |
| LDL | 1.96 (1.23-3.20) | 0.9(0.48-4.20) | 89402.00 | 0.069 |
| HDL | 1.4 (1.1-1.8) | 1.07(0.80-1.30) | 52475.00 | <0.001 |
| Triglycerides | 1.15 (0.82-1.54) | 0.99(0.68-1.47) | 94871.00 | 0.643 |

### Predictive Model Performance and Selection

Eight ML models were evaluated using 5-fold cross-validation. The RF model achieved the highest mean accuracy of 95.18+/-1.33%, PPV of 92.80+/-4.39%, sensitivity of 87.78+/-3.91%, F1-score of 90.10+/-2.64%, and AU-ROC of 0.97. AdaBoost followed with 94.69+/-1.14% accuracy, 89.93+/-3.81% PPV, 88.98+/-3.83% sensitivity, and 89.34+/-2.26% F1-score. Naïve Bayes performed the lowest, with 85.73+/-2.21% accuracy, and 67.01+/-6.83% F1-score (Table 2).

**Table 2:**
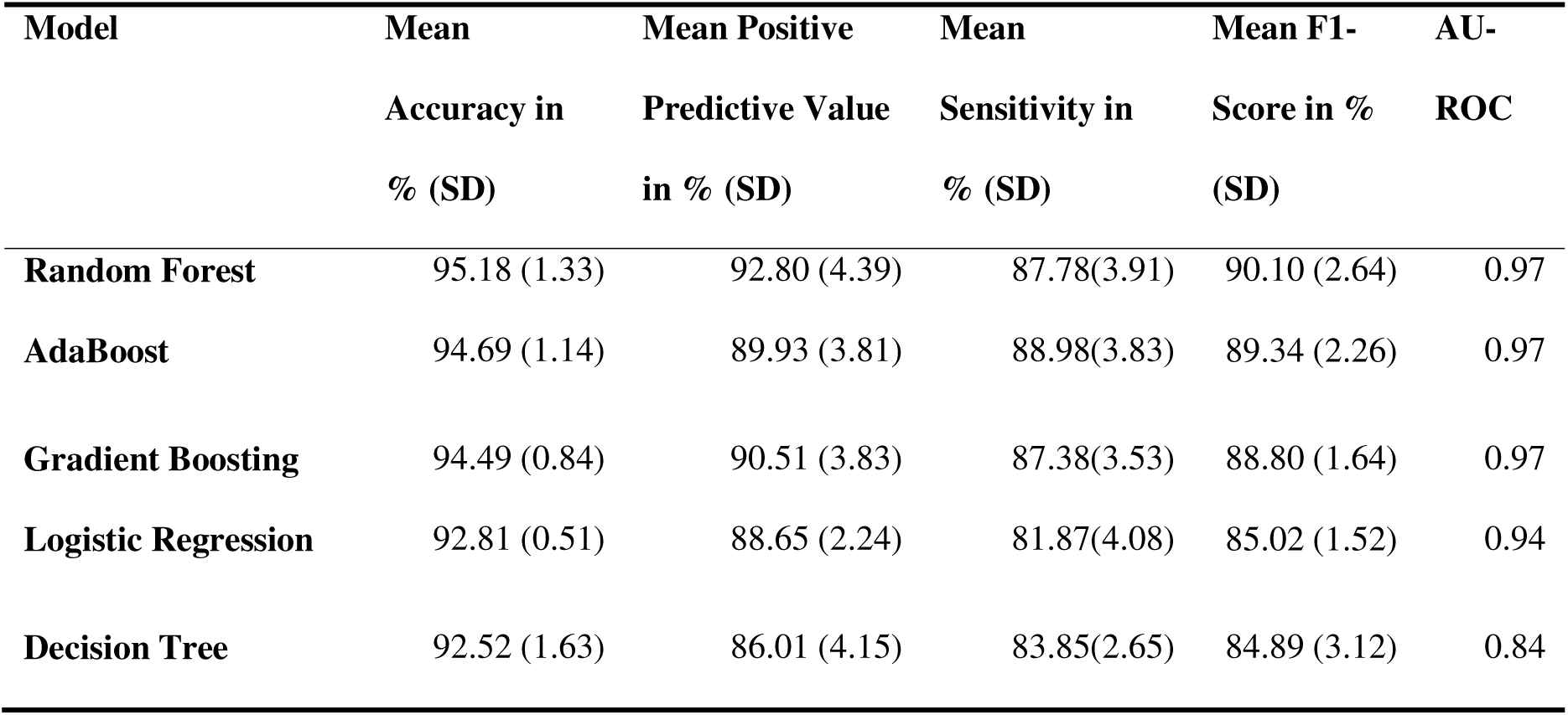
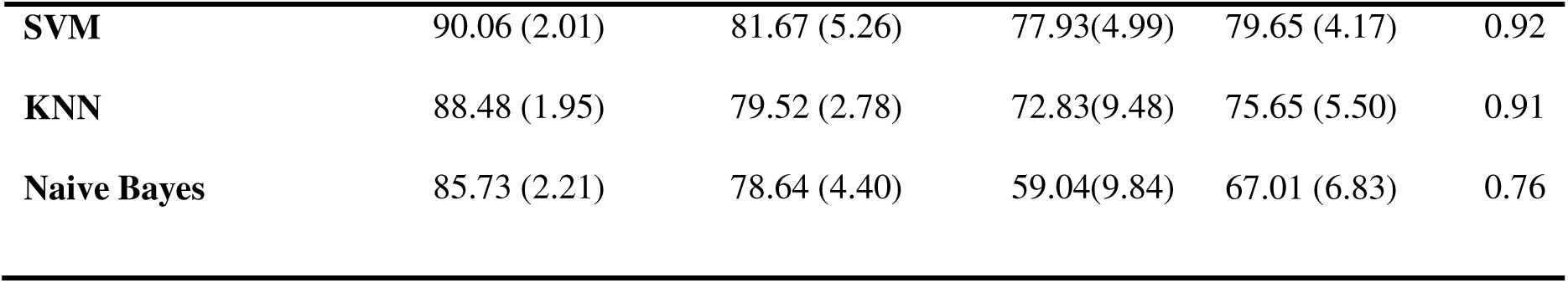
Performance metrics of eight machine learning models for stroke risk prediction.

| Model | Mean<br>Accuracy in<br>% (SD) | Mean Positive<br>Predictive Value<br>in % (SD) | Mean<br>Sensitivity in<br>% (SD) | Mean F1-Score in %<br>(SD) | AU-ROC |
| --- | --- | --- | --- | --- | --- |
| Random Forest | 95.18 (1.33) | 92.80 (4.39) | 87.78(3.91) | 90.10 (2.64) | 0.97 |
| AdaBoost | 94.69 (1.14) | 89.93 (3.81) | 88.98(3.83) | 89.34 (2.26) | 0.97 |
| Gradient Boosting | 94.49 (0.84) | 90.51 (3.83) | 87.38(3.53) | 88.80 (1.64) | 0.97 |
| Logistic Regression | 92.81 (0.51) | 88.65 (2.24) | 81.87(4.08) | 85.02 (1.52) | 0.94 |
| Decision Tree | 92.52 (1.63) | 86.01 (4.15) | 83.85(2.65) | 84.89 (3.12) | 0.84 |
| **SVM** | 90.06 (2.01) | 81.67 (5.26) | 77.93(4.99) | 79.65 (4.17) | 0.92 |
| **KNN** | 88.48 (1.95) | 79.52 (2.78) | 72.83(9.48) | 75.65 (5.50) | 0.91 |
| **Naive Bayes** | 85.73 (2.21) | 78.64 (4.40) | 59.04(9.84) | 67.01 (6.83) | 0.76 |

Table 3 shows the McNemar’s test contingency table comparing RF and AdaBoost classification performance. Of 1,016 predictions, both models correctly classified 945 cases and misclassified 37. RF correctly classified 25 cases misclassified by AdaBoost with a significant difference (p=0.01).

**Table 3:**
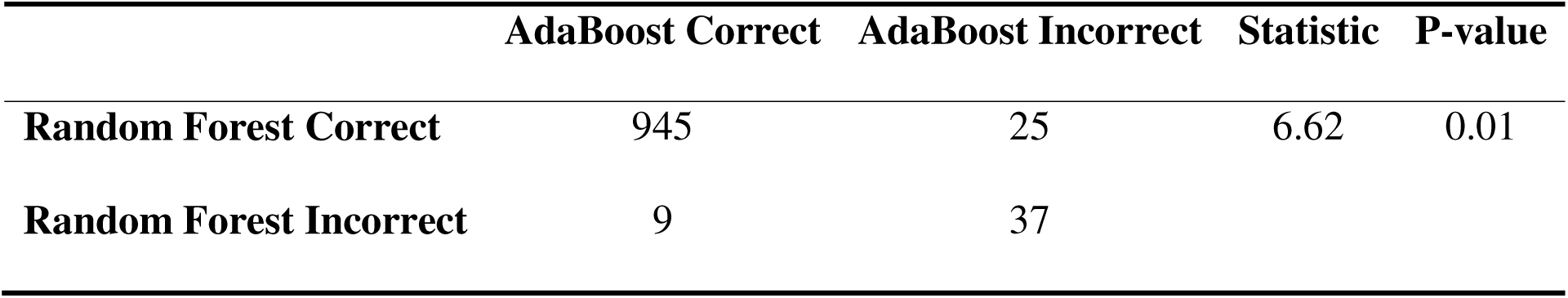
Contingency table for McNemar’s test comparing the classification performance of Random Forest and AdaBoost models (n=1016).

|  | **AdaBoost Correct** | **AdaBoost Incorrect** | **Statistic** | **P-value** |
| --- | --- | --- | --- | --- |
| **Random Forest Correct** | 945 | 25 | 6.62 | 0.01 |
| **Random Forest Incorrect** | 9 | 37 |  |  |

### Predictors Influencing the Random Forest Model Predictions

Feature importance analysis of the RF model ranked age, systolic blood pressure, total cholesterol, and waist-hip ratio as the top predictors of stroke risk, with pulse rate as the least influential (Figure 1).

**Fig 1:**
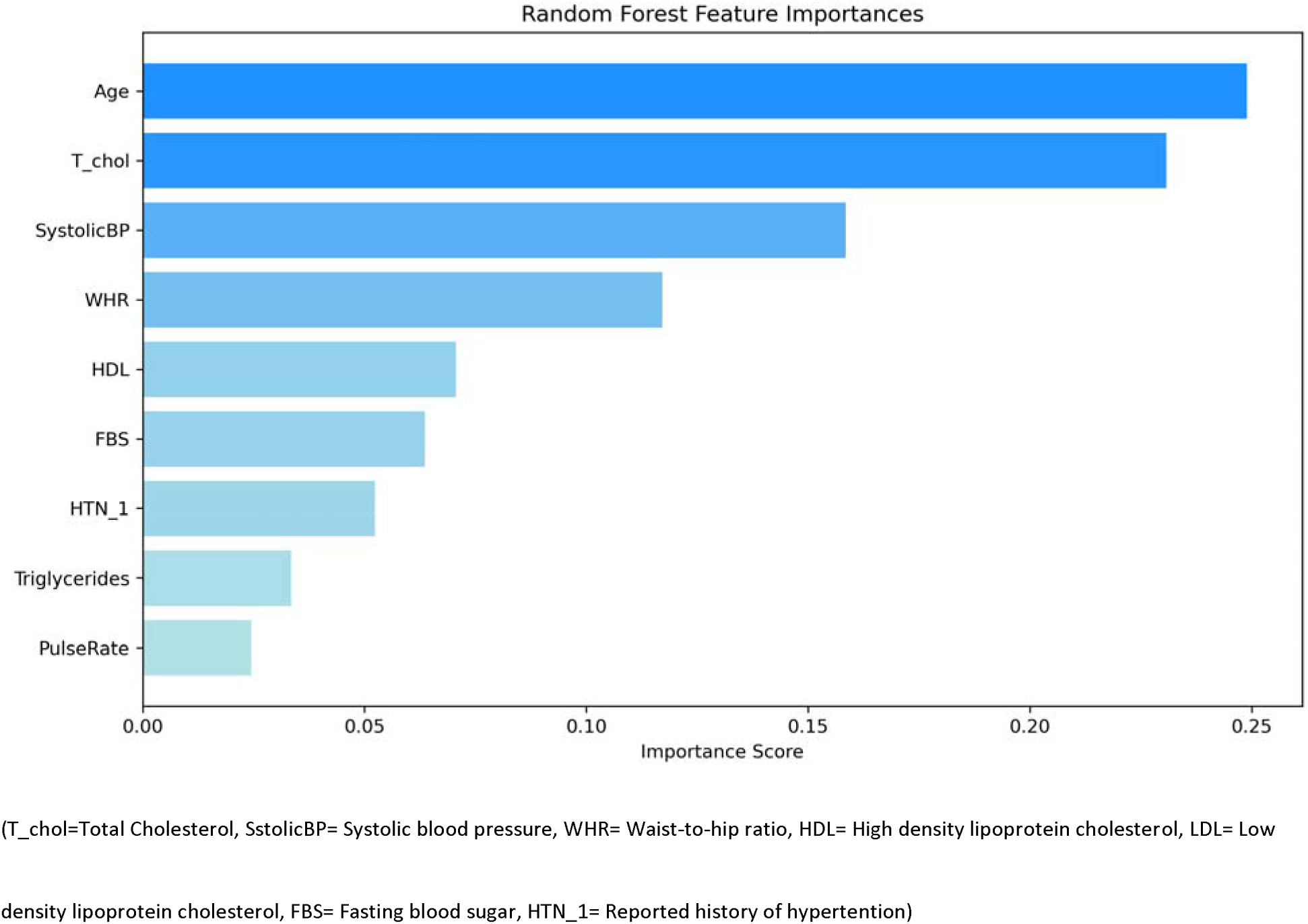
Bar plot showing feature importance scores from the Random Forest model for stroke risk prediction.

### Misclassification Patterns

The RF model, evaluated using 5-fold cross-validation, misclassified 21(8.3%) stroke cases as non-stroke (false negatives) and 25(3.3%) non-stroke cases as stroke (false positives), as shown in the classification matrix (Figure 2).

**Fig 2:**
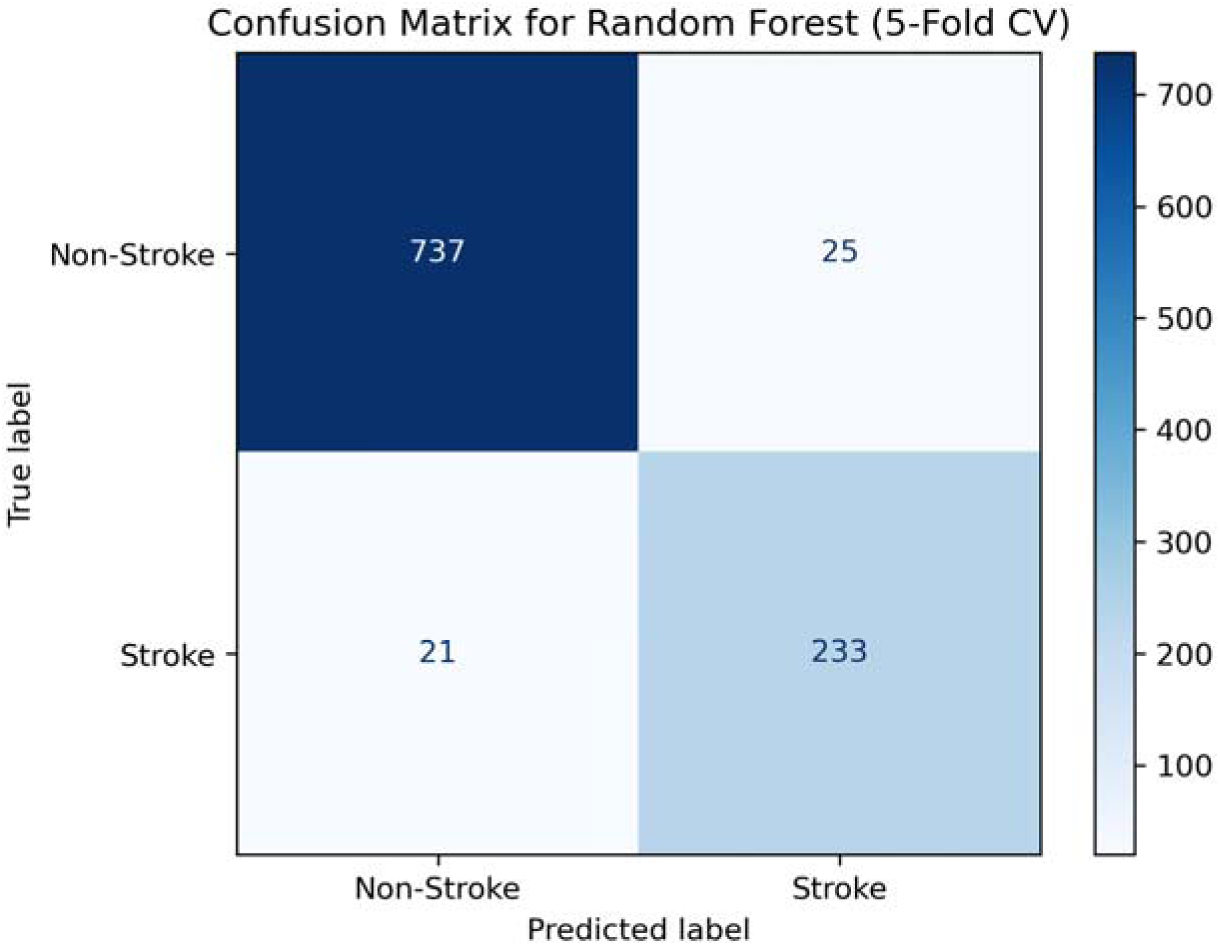
Classification matrix showing the aggregated predictions of the Random Forest model across 5-fold cross-validation (n=1016).

Figure 3 displays the distribution of false positives and false negatives across a range of decision thresholds. As the threshold increases, the number of false positives declines steadily, while the number of false negatives rises sharply. At the default threshold of 0.5, the plot shows a relative balance between false positives (25) and false negatives (21).

**Fig 3:**
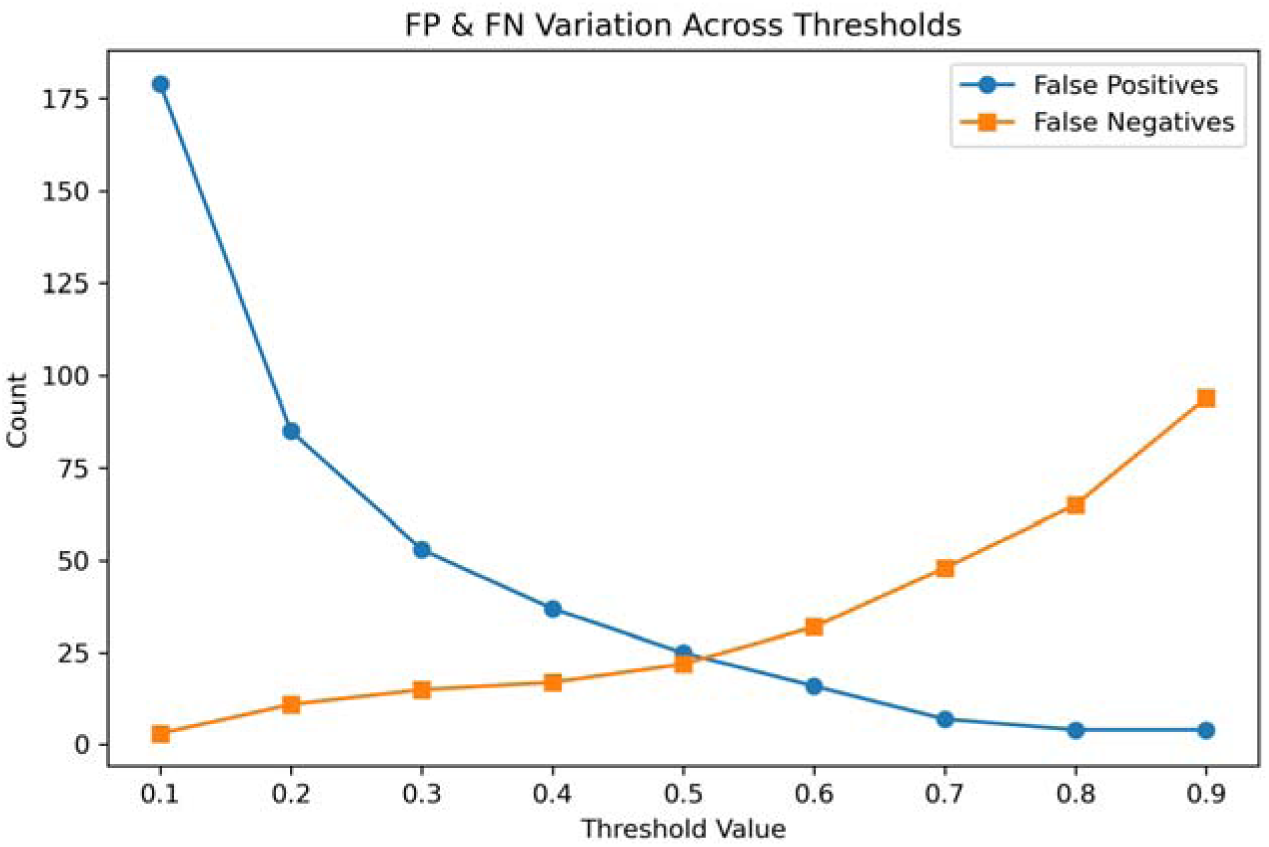
Line graph showing misclassification trends across varying decision thresholds of the Random Forest model for stroke risk prediction.

False Negatives had lower total cholesterol (3.67 mmol/L, IQR: 3.13-4.58) and systolic blood pressure (131 mmHg, IQR: 103-148) than true positive cases, while false positives had higher values (4.83 mmol/L, IQR: 3.9-5.6; 145 mmHg, IQR: 121-161) than true negatives, as detailed in Table 4.

**Table 4:**
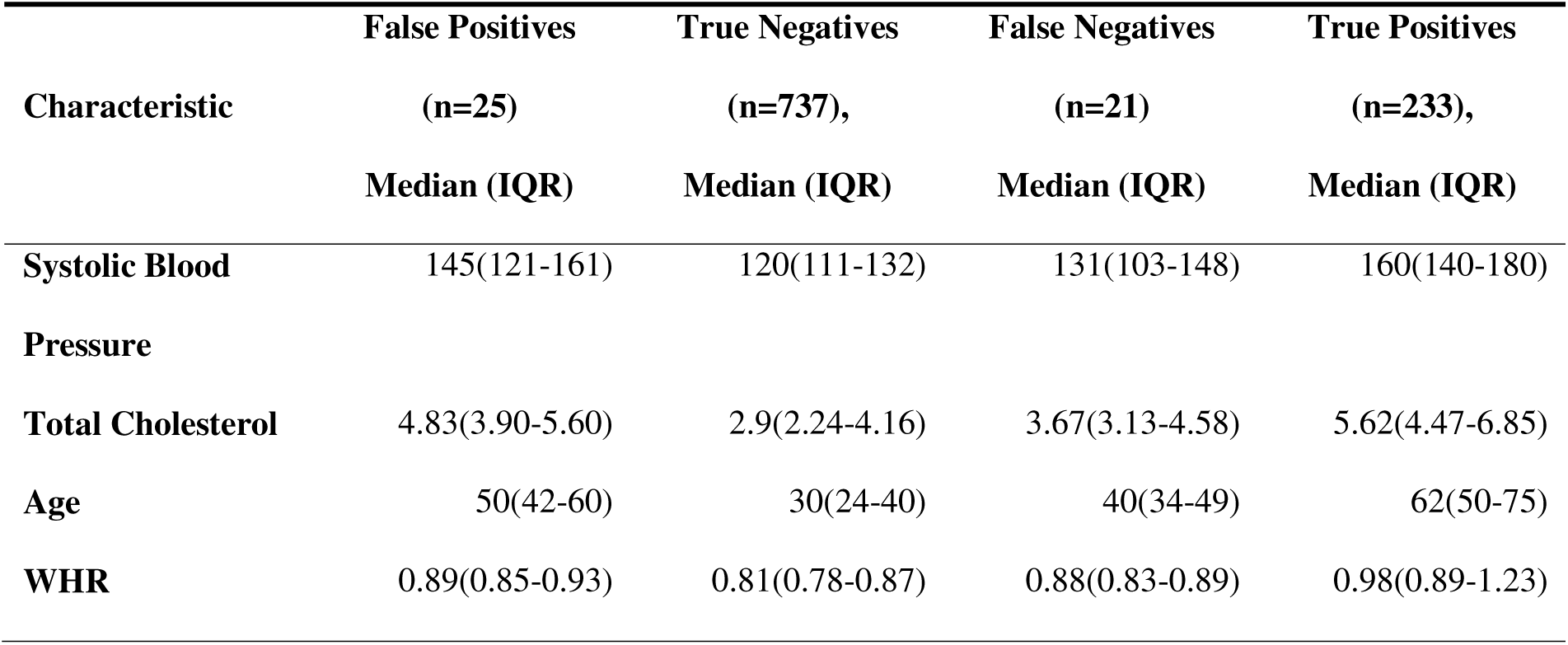
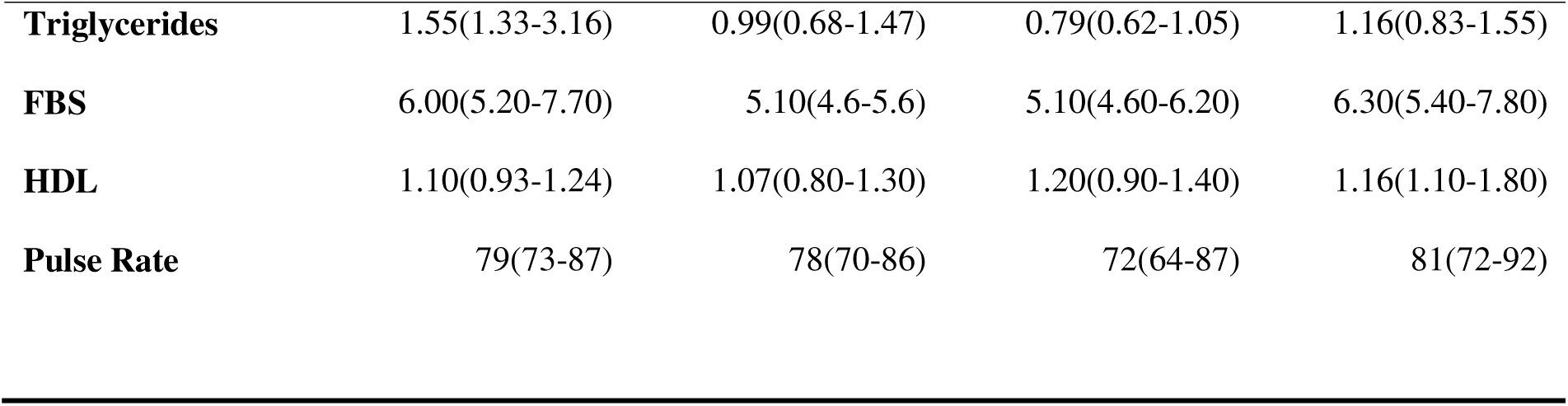
Clinical characteristics of misclassified and correctly classified cases by the Random Forest model.

|  | False Positives<br>(n=25)<br>Median (IQR) | True Negatives<br>(n=737),<br>Median (IQR) | False Negatives<br>(n=21)<br>Median (IQR) | True Positives<br>(n=233),<br>Median (IQR) |
| --- | --- | --- | --- | --- |
| **Systolic Blood Pressure** | 145(121-161) | 120(111-132) | 131(103-148) | 160(140-180) |
| **Total Cholesterol** | 4.83(3.90-5.60) | 2.9(2.24-4.16) | 3.67(3.13-4.58) | 5.62(4.47-6.85) |
| **Age** | 50(42-60) | 30(24-40) | 40(34-49) | 62(50-75) |
| **WHR** | 0.89(0.85-0.93) | 0.81(0.78-0.87) | 0.88(0.83-0.89) | 0.98(0.89-1.23) |
| **Triglycerides** | 1.55(1.33-3.16) | 0.99(0.68-1.47) | 0.79(0.62-1.05) | 1.16(0.83-1.55) |
| **FBS** | 6.00(5.20-7.70) | 5.10(4.6-5.6) | 5.10(4.60-6.20) | 6.30(5.40-7.80) |
| **HDL** | 1.10(0.93-1.24) | 1.07(0.80-1.30) | 1.20(0.90-1.40) | 1.16(1.10-1.80) |
| **Pulse Rate** | 79(73-87) | 78(70-86) | 72(64-87) | 81(72-92) |

## Discussion

The RF model outperformed alternatives such as AdaBoost in 5-fold cross-validation, achieving a mean F1-score of 90.1%. This superior performance is likely due to RF’s ensemble learning strategy, which averages multiple decision trees to reduce overfitting and enhance robustness to noisy or imbalanced data (17). These findings are consistent with prior stroke prediction studies that highlight RF’s adaptability to diverse data patterns. For example, Shobayo et al. (2023) reported an RF model with 94% F1-score and 94% accuracy in Bangladesh, while a Nigerian study found 93% accuracy (18). In contrast, some studies have identified other algorithms as superior. A study in India reported XGBoost achieving a 96.8% accuracy and 98% AU-ROC, surpassing RF’s 94.2% accuracy, while Li et al. (2020) in China found SVM yielding 93.5% accuracy and 96% AU-ROC, outperforming RF (91.8%). Differences in top-performing algorithms across studies likely arise from dataset-specific traits, such as feature dimensionality or class imbalance, with this study’s comprehensive clinical features and SMOTE application enhancing RF’s robustness to noise (17). The RF model’s strong performance suggests potential for stroke risk stratification in Uganda, where diagnostic resources are limited and stroke mortality is high (4).

However, this strong performance may be partly inflated by confounding from the unmatched case-control design, necessitating external validation to confirm generalizability across diverse Ugandan populations.

The RF model identified age, systolic blood pressure, total cholesterol, and waist-hip ratio as top predictors of stroke risk, with additional contributions from fasting blood sugar, HDL cholesterol, triglycerides, and pulse rate.

Age ranked as the most influential predictor in the RF model, reflecting its critical role in stroke risk stratification due to its ability to capture cumulative vascular aging. This aligns with an African systematic review’s 3-4% annual risk increase in African populations, where each decade roughly doubles stroke odds (19) and the Framingham Study which reported an age effect of 66% and 93% likelihood of stroke per decade for men and women respectively (20). This highlights age’s biological role in increasing arterial stiffness and cumulative risk factor exposure, as aging accelerates endothelial dysfunction and atherosclerosis, making older individuals prime candidates for stroke (21). The role of age in this study, however, may be overestimated, as stroke cases had a significantly higher median age (60 years) compared to community controls (30 years). This gap is partly reflective of Uganda’s youthful community population, where the national median age is approximately 16.9 years. The resulting age disparity may have amplified age’s predictive weight in the RF model, elevating its influence as a top-ranked feature. The RF model’s emphasis on age demonstrates its strength in prioritizing older Ugandans for screening, particularly at referral centers like Mulago hospital, where severe strokes predominate.

Systolic blood pressure was a leading predictor in the RF model, capturing hypertension’s pivotal role in stroke pathogenesis. This is consistent with the INTERSTROKE study which found a 0.4-0.6% risk increase per mmHg in African cohorts, where hypertension almost triples overall stroke risk (8). This is possibly because elevated systolic blood pressure increases stroke risk by causing chronic endothelial injury and accelerating atherosclerosis, leading to arterial stiffening and plaque formation that predispose to ischemic stroke, while high pressure can also rupture weakened cerebral vessels, triggering hemorrhagic stroke (22). RF’s prioritization of systolic blood pressure highlights its power to detect clinically measurable risk factors, supporting routine blood pressure monitoring in Uganda’s primary care to reduce stroke burden.

Total cholesterol emerged as a strong predictor in the RF model, highlighting its atherogenic contribution to stroke risk. This aligns with the 23% increase reported among individuals of African ancestry in the United Kingdom (23), and findings from the SIREN study in West Africa, where dyslipidemia was associated with more than a fivefold increase in stroke risk (24). The significance observed across studies is linked to the role of cholesterol in promoting atherosclerotic plaque formation within cerebral arteries, thereby increasing the risk of ischemic stroke (25). The RF model’s reliance on total cholesterol highlights its capacity to detect dyslipidemia using routine lipid profiles; an accessible and cost-effective screening approach suitable for resource-limited settings.

WHR, a marker of visceral obesity, emerged as a key predictor in the RF model, reflecting the metabolic contribution to stroke risk. This aligns with findings from a Ugandan study conducted at Mulago Hospital, where young adults with higher WHR were over 11 times more likely to experience stroke, emphasizing the strong local relevance of central obesity as a risk factor (26). This is because visceral fat promotes chronic inflammation, insulin resistance, and dyslipidemia, accelerating atherosclerosis, thus increasing risk of cerebrovascular events (27). In a broader African context, the INTERSTROKE study reported a 69% increased stroke risk per unit rise in WHR among African populations (28). The variation in effect estimates across these studies likely reflects differences in age distribution, baseline metabolic health, and study design, but collectively reinforces the role of central obesity as a significant and modifiable stroke risk factor. The role of WHR in this study may be inflated by age disparity between cases and controls, as older individuals; who may have higher WHR (29) were overrepresented among stroke patients, suggesting possible positive confounding by age. RF’s emphasis on waist-hip ratio highlights its power to detect anthropometric risk factors, supporting prioritization of obesity management in Uganda.

HDL cholesterol was a key predictor in the RF model. Unexpectedly, stroke cases had higher median HDL levels than controls, contrasting with HDL’s typical protective role against stroke via reverse cholesterol transport. Typically, HDL cholesterol is believed to reduce stroke risk by promoting reverse cholesterol transport and inhibiting atherosclerotic plaque formation (30). For instance, the INTERSTROKE study observed a 25% protective effect of high HDL levels in their African cohort (31). However, counterintuitive findings similar to this study’s, have been reported in other settings. A prospective longitudinal study in China found a U-shaped relationship between HDL and stroke, with individuals having HDL ≥ 2.05 mmol/L showing up to an 85% increased risk of ischemic stroke compared to those in the moderate HDL range (32). These findings support the possibility that elevated HDL may not universally offer protection and could indicate increased stroke risk in some populations. This is possibly because HDL functionality, rather than concentration alone, determines its protective effect (30,33). In the presence of chronic inflammation or oxidative stress, HDL can become dysfunctional, losing its anti-inflammatory and antioxidant properties and potentially becoming pro-atherogenic (30). Additionally, regional differences in genetics, comorbidities, and environmental exposures may influence HDL behavior across populations (33). The reliance of the RF model on HDL cholesterol as a key predictor highlights its ability to detect complex, non-linear relationships; potentially reflecting interactions with other lipids such as total cholesterol. It is also possible that elevated HDL levels in stroke cases reflect a generalized increase in lipid levels, including total cholesterol, rather than HDL exerting an independent effect. However, RF feature importance does not indicate the direction of HDL’s effect, highlighting the need for future research to incorporate more interpretable modeling approaches, such as hybrid models, that can offer both predictive power and explanatory clarity. Additionally, future studies should investigate the functional quality of HDL and its longitudinal dynamics to better understand its role in stroke risk stratification and to enhance the clinical utility of RF-based prediction tools. In high-stroke-burden settings like Mulago hospital, routine lipid screening remains essential.

Fasting blood sugar emerged as a key predictor in the RF model, highlighting the role of impaired glucose regulation in stroke pathophysiology. This aligns with the well-established link between DM and vascular damage, which accelerates atherosclerosis and increases cerebrovascular event risk. For instance, the SIREN study in West Africa reported that higher fasting blood sugar was associated with a 4.29-fold increased risk of stroke, demonstrating the substantial impact of hyperglycemia in African settings (24). However, Namaganda et al. (2022) reported no significant association between fasting blood sugar and stroke in a study focused on young adults at Mulago Hospital. This discrepancy could reflect age-related differences in DM prevalence and cumulative vascular injury, suggesting that the impact of hyperglycemia on stroke risk may be more pronounced in older populations. The inclusion of fasting blood sugar as a key predictor in the RF model illustrates the model’s ability to identify clinically relevant metabolic risk factors, supporting the importance of glucose monitoring and DM control, particularly in rapidly urbanizing regions like Wakiso District.

History of hypertension was a key predictor in the RF model, considered separately from systolic blood pressure, as not all diagnosed individuals presented with elevated readings at the time of data collection. In this study, 59.1% of stroke cases had a documented history of hypertension, compared to only 5.4% of controls, highlighting its pivotal role in stroke pathogenesis through mechanisms such as endothelial dysfunction, arterial remodeling, and vascular stiffening (22). These findings are broadly consistent with the INTERSTROKE study, which reported hypertension in 55% of African stroke cases and 22% of controls (8). O’Donnell et al. (2016) estimated a 47.9% increased stroke likelihood among hypertensive individuals in Africa, whereas the SIREN study reported a substantially higher likelihood 90.8% in Nigeria and a comparatively lower 19.36% in Ghana (35). These variations likely reflect differences in hypertension prevalence, awareness, treatment access, and genetic predispositions across regions. That the RF model recognized history of hypertension as a significant predictor in this study emphasizes the importance of context-specific modeling and highlights the need for tailored hypertension management strategies in Uganda, especially in rapidly urbanizing settings like Wakiso.

Though modest, triglycerides had a role in the RF model predictions in this study. A systematic review of nine prospective cohort studies found a 10% increased risk of stroke per one standard deviation increase in triglycerides (36). This is because triglycerides contribute to stroke risk by promoting endothelial dysfunction, inflammation, and the formation of atherogenic remnant particles, which accelerate arterial plaque development (36,37). However, some studies in African populations found no significant association between triglycerides and stroke after multivariate adjustment. For instance, a retrospective cross-sectional study in Nigeria (38) and a retrospective cohort study in Ghana (24) reported non-significant findings, suggesting that this relationship may vary by region, population structure, or comorbid profile. The RF model identified triglycerides as a useful predictor, likely due to its ability to capture complex, non-linear interactions (17,39) and interactions with other metabolic variables. RF may recognize subtler patterns where elevated triglycerides contribute to stroke risk in combination with other features like blood pressure, waist-hip ratio, or glucose levels; relationships linear models may overlook. Further prospective studies are warranted to clarify the contextual role of triglycerides in stroke pathogenesis across diverse African populations, and to evaluate their utility in locally adaptable risk prediction tools.

Pulse rate emerged as the least influential predictor in the RF model. Pulse rate has not widely been referred to as a well-established predictor of stroke in most studies, unlike the REGARDS study West Africa, which reported a 10% increased stroke risk per 10 beats per minute rise in resting pulse rate (40). This discrepancy may reflect population differences, measurement timing, or confounding by comorbidities. In this study, pulse rate may function more as an indirect marker, reflecting physiological stress or autonomic imbalance rather than acting as a primary causal factor in stroke pathogenesis. It is possible that some elevated pulse rate readings were a response to stroke rather than a true pre-event predictor. Moreover, elevated pulse rate is often a surrogate for underlying cardiovascular conditions such as autonomic dysfunction, systemic inflammation, or poorly controlled hypertension, factors that are themselves directly associated with stroke risk. The RF model’s inclusion of pulse rate, despite its low feature importance score, illustrates the model’s sensitivity to subtle or non-linear relationships. While pulse rate may not be a strong standalone predictor, its presence in the model suggests potential utility in composite risk estimation. Continued cardiovascular monitoring, including pulse rate, remains relevant within a broader predictive framework, particularly in resource-limited settings where advanced diagnostics may be limited.

The RF model’s feature selection excluded history of smoking and harmful alcohol use, despite their established roles in stroke pathogenesis. History of smoking, though a well-documented stroke risk factor in global literature, was excluded by the RFECV (hence not included in the RF model’s feature list). This contrasts with prior evidence such as the INTERSTROKE study, which reported a 67% increased odds of stroke among smokers, with a 12.4% population-attributable risk in African cohorts (8). The significance of smoking in other previous research reflects its well established biological mechanisms in promoting endothelial dysfunction, accelerating atherosclerosis, and inducing pro-thrombotic states (41), all of which elevate ischemic stroke risk. The discrepancy in findings may be due to the low prevalence of reported smoking in this study with no current smokers and 6.54% former smokers (Table 1), which weakens its statistical signal. Underreporting bias from retrospective medical records may have further limited the accuracy and variability of the smoking variable, reducing its contribution in this study. Given the strong and consistent associations reported in multicenter studies, caution is warranted when interpreting the effect of smoking in this study. These findings highlight the need for locally conducted longitudinal studies to more accurately determine the role of smoking in the stroke causation pathway within Ugandan populations.

Harmful alcohol use was also excluded by RF’s feature selection. This is contradictory to alcohol’s known association with stroke based on available literature. For instance, INTERSTROKE’s 60% risk increase for heavy alcohol use in African cohorts (42). A Nigerian study found a stronger effect, with heavy alcohol use increasing stroke risk by 100% (43), possibly due to higher prevalence or cultural differences in alcohol consumption patterns. This study’s cases reported harmful alcohol use in only 13.7% (Table 1), lower than INTERSTROKE’s African cohort (25%) (42), likely attenuating its signal. Therefore, RF’s exclusion of harmful alcohol use may be attributed to its low observed prevalence, as the model prioritizes features with higher variability and predictive contribution. However, given that Uganda ranks among the countries with the highest alcohol consumption (44), the prevalence in this study is likely underestimated due to underreporting bias. These findings should therefore, be interpreted with caution, and further research particularly using prospectively collected data, is needed to clarify the role of alcohol consumption in stroke risk stratification within Ugandan and similar populations.

Despite the RF model’s robust performance, misclassification patterns highlight areas for refinement, with 21 stroke cases (8.3%) misclassified as non-stroke (false negatives, FN) and 25 non-stroke cases (3.3%) as stroke (false positives, FP) (Figure 2). These errors, driven by overlaps in risk profiles and data limitations, highlight the need for calibration to enhance RF’s utility in Uganda’s high-mortality stroke context.

False negatives often had risk profiles resembling true negatives. This suggests that the primary cause of stroke may be due to unaccounted for risk factors like atrial fibrillation, genetic predisposition, or transient risks (e.g., infections) likely contributed (45–48). Data quality issues, such as class imbalance, despite use of SMOTE, and RFECV’s exclusion of variables like harmful alcohol use may have further limited RF’s sensitivity. Missing these cases risks delaying interventions in Uganda, where stroke mortality is high (4).

False positives exhibited elevated risk profiles, with higher systolic blood pressure and total cholesterol than true negatives, resembling true positives (Table 4). This suggests that RF can identify individuals with heightened stroke risk who have not yet experienced an event, reinforcing its proactive role in identifying at-risk individuals for prevention in Uganda’s resource-limited setting. While false positives may indicate early risk detection, misclassifying low-risk individuals as high-risk warrants caution, as it can lead to unnecessary anxiety, repeat testing, and potential overuse of limited healthcare resources.

The observed false positive and false negative rates may have been over- or underestimated due to threshold-related effects, as their counts shift notably across varying decision thresholds. As illustrated in Figure 3, the number of false positives declines steadily with increasing thresholds, while false negatives rise sharply; intersecting near the default threshold of 0.5. This reflects a point of maximum classification uncertainty, where the model is equally prone to both types of error. These trends directly influence sensitivity and specificity. Wallace et al., 2023 highlighted similar threshold issues in RF stroke models, recommending calibration to balance sensitivity and specificity, with 0.33 as their optimal predicted probability threshold. Adjusting the classification threshold by lowering the threshold to enhance sensitivity could reduce FN cases but could strain Uganda’s limited resources (12) by increasing diagnostic and follow-up demands. Conversely, raising it may reduce false positives but risk missing subtle cases (12). This highlights the threshold-dependent nature of the Random Forest model’s performance and the need for careful calibration based on clinical priorities. Introducing an intermediate-risk category or applying post-hoc calibration techniques (50) could help resolve ambiguity in borderline predictions and support more balanced clinical decision-making. Use of longitudinal data to track predictor changes could also enhance RF’s precision for stroke prevention in Uganda’s high-burden setting.

### Study Limitations

The unmatched case-control design introduces confounding particularly due to age disparity, which may bias predictions. The ‘cohort effect’ and potential data quality inconsistencies arising from heterogeneous retrospective sources collected across different years may have introduced differential misclassification bias. This was particularly likely for self-reported variables such as smoking and alcohol use, despite mitigation efforts through consistent PI oversight. The use of secondary data limited the availability of certain variables, including key potential confounders like physical activity and dietary habits, and introduced challenges related to missing data. Furthermore, this study modeled stroke as a single composite outcome without distinguishing between ischemic and hemorrhagic subtypes, which may obscure subtype specific risk factor associations given the differing pathophysiological mechanisms and risk profiles of these stroke types. Limitation of recall, reporting bias, and the use of surrogate respondents for severely ill stroke patients; who made up the majority, may have led to underestimation of self-reported variables. This could have, in turn, affected the relevance and selection of these variables in the RF model. The trade-off between RF’s complexity and interpretability restricted clinical actionable insights, such as direction of predictor effects. The lack of longitudinal validation and unmeasured treatment effects may further limit the model’s real-world accuracy and applicability. Imbalanced data may have biased RF toward non-stroke predictions, reducing sensitivity despite SMOTE application. The focus on MNRH and Wakiso District limits generalizability to other Ugandan regions, where stroke risk profiles may differ.

## Conclusions

The Random Forest model outperformed other models, correctly classifying approximately 95 of every 100 participants as stroke or non-stroke cases. Age, systolic blood pressure, total cholesterol, and waist-hip ratio were the top predictors influencing the RF model predictions. Misclassification pattern assessment revealed overlapping risk profiles and prediction dynamics threshold as the key challenges for the RF model performance.

The Ministry of Health should consider integrating ML-based tools like the RF model into clinical workflows for early stroke risk detection. Strengthening routine documentation of key risk factors in primary care will improve both prevention and model reliability. Future studies should validate the model in diverse populations and explore hybrid approaches that balance accuracy with interpretability.

## Data Availability

Data is available upon request from the corresponding author

## Non-standard Abbreviations and Acronyms

AU-ROC: Area Under Receiver Operator characteristics Curve
EPV: Events Per Variable
WHR: Waist-to-hip ratio
k-NN: K-Nearest Neighbors
LMICs: Low and Middle Income Countries
ML: Machine Learning
PI: Principal Investigator
RF: Random Forest
RFECV: Recursive Feature Elimination with Cross-Validation

## Declarations

### Ethics approval and consent to participate

Permission to conduct the study was sought from the Clinical Epidemiology Unit, Makerere University. Ethical approval for use of secondary data, was sought from the Makerere University School of Medicine Research and Ethics Committee (Mak-SOMREC-1200). Informed consent for the use of secondary data was deemed obtained through the primary studies, and Mak-SOMREC was requested to waive participant consent for this study.

### Consent for publication

Not applicable.

### Authors’ Contributions

DM: conceptualization, protocol writing, ethical approval, database design, data collection oversight, analysis, manuscript drafting.

BW, HK: Methods, protocol and manuscript review

DW, SN, SO, HU: data collection, manuscript writing and review.

JK: Conceptualization, methodological oversight, protocol and manuscript review

JN, MK-M, RB: supervision, methods, manuscript review. All authors approved the final manuscript.

## Acknowledgements

Prof. Joan N. Kalyango is appreciated for her invaluable mentorship, and Dr. Jane Nakibuuka (PI of the primary studies), Dr. Martin Kaddu-Mukasa, and Prof. Rehema Baguma for their expert supervision and critical guidance; also grateful to the research assistants and colleagues whose support made this work possible.

